# Screening sham mirror-therapy materials using fNIRS: protocol for an acute within-participant randomised crossover mechanistic study

**DOI:** 10.64898/2026.08.23.26361088

**Authors:** Tangzhu Yang, Yuezhu Wang, Siyuan Wei, Dingqun Bai

## Abstract

**Introduction:** Selecting an appropriate sham control is a key challenge in trials of mirror therapy—specifically paradigms using mirror visual feedback (MVF), because visually similar control conditions may still elicit mirror-related cortical responses. This protocol describes an acute mechanistic, within-participant fNIRS screening study designed to identify the sham mirror-therapy material condition with the most “neutral” neural signature relative to true MVF during a single exposure.

**Methods and analysis:** This is a single-centre, within-participant, randomised crossover study conducted at Wuhan Wuchang Hospital (Wuhan, China). Healthy adults aged 18–35 years will complete four conditions in one visit: C1 true MVF and three prespecified sham-material conditions (C2–C4), with condition order counterbalanced using a Latin-square schedule. fNIRS will be acquired during a standardised grasping task. Online acquisition-time quality control (SCI and CV thresholds) will be applied with prespecified re-acquisition rules. The primary outcome is ROI-level task-evoked change in oxygenated haemoglobin (ΔHbO) within prespecified ROIs (PMC and SM1/M1), estimated primarily using GLM-derived β estimates. Condition effects will be analysed using linear mixed-effects models with prespecified contrasts and Holm multiplicity adjustment to rank sham conditions by a prespecified neutrality decision rule.

**Ethics and dissemination:** Ethics approval was obtained from the Ethics Committee of Wuchang Hospital Affiliated to Wuhan University of Science and Technology (Approval No.: 2025-112-01). Findings will be disseminated through publication of this protocol manuscript and a subsequent results manuscript, with key supplementary materials provided as online appendices/supplements as required by the target journal.

**Trial registration number:** Chinese Clinical Trial Registry (ChiCTR2600116634).

**Strengths and limitations of this study:** **Within-participant randomised crossover design** reduces between-participant variability and is well suited for acute mechanistic screening of sham conditions.

**Prespecified sham conditions and neutral-ranking decision rule** (including prespecified ROIs, contrasts, and Holm multiplicity control) help limit analytic flexibility and support transparent interpretation.

**Operational reproducibility safeguards** are specified, including standardised task timing/instructions, acquisition-time QC thresholds (SCI/CV) with re-acquisition rules, and frozen channel-to-ROI mapping and material-definition records in the Supplementary materials.

**Single-centre, healthy-participant, single-session paradigm** may limit generalisability to clinical stroke populations and to longer-term therapeutic effects.

**Blinding may be imperfect** because perceptual differences between materials can affect expectancy/attention; blinding assessment is included but residual bias is possible.

**fNIRS is susceptible to motion/scalp-coupling variability and physiological noise**; despite prespecified QC and preprocessing, residual artefacts may remain and can reduce sensitivity.

## Introduction

Mirror therapy—specifically paradigms using mirror visual feedback (MVF)—is widely used in neurorehabilitation for improving upper-limb motor function after stroke and other neurological conditions ^[1]^. However, in randomised controlled trials (RCTs), selecting an appropriate control condition remains challenging because visually or contextually similar sham conditions may still engage action-observation and motor networks—particularly premotor and sensorimotor regions (e.g., PMC and SM1/M1)—thereby diluting contrast and complicating interpretation of MVF-specific effects ^[2-4]^.

Importantly, the sham/control conditions used across prior MVF studies are highly heterogeneous (e.g., opaque occluders, transparent panels, non-reflective materials, or bilateral synchronous movement paradigms), which reduces cross-study comparability and makes it difficult to interpret whether observed effects are MVF-specific or driven by non-specific contextual/visual stimulation ^[2, 5]^ . Moreover, few studies have directly compared candidate sham conditions at the neural level (i.e., neutrality of motor-network activation) or provided a reproducible, versioned record that freezes material IDs/names and key implementation specifications^[5-7]^ . These gaps motivate a screening study with prespecified candidate conditions and a prespecified fNIRS-based neutrality ranking framework (ROIs, contrasts, and multiplicity control), to minimise design flexibility and provide an evidence-based rationale for sham selection prior to subsequent confirmatory RCTs^[8, 9]^.

An optimal sham condition should therefore match the non-specific aspects of the intervention context (e.g., posture, attention demands, task timing, and experimenter–participant interaction) while minimising MVF-specific visual feedback that could trigger mirror-related neural activation ^[2, 5]^. Reporting standards for nonpharmacologic trials and intervention descriptions further underscore the need to define and standardise control conditions with sufficient operational detail to enable replication and to reduce analytic and interpretive flexibility^[2, 5, 10, 11]^.

Functional near-infrared spectroscopy (fNIRS) provides a practical, non-invasive approach for quantifying task-evoked cortical haemodynamic responses in regions implicated in action observation and motor control (e.g., premotor and sensorimotor cortices) ^[12-15]^ . Accordingly, fNIRS can be used as an acute mechanistic screening tool to compare candidate sham materials under highly standardised conditions and to identify the option with the most neutral neural signature relative to true MVF ^[16]^.

From a trial-design perspective, conducting this mechanistic screening *before* a confirmatory RCT helps avoid embedding an inadequately specified or biologically active sham condition into a costly efficacy trial ^[2, 5]^ . Here, neutral is operationally defined primarily as the sham condition that produces the smallest absolute task-evoked fNIRS response in prespecified motor-network ROIs among the sham conditions. Comparisons with true MVF will be interpreted by prioritising smaller absolute motor-network activation in the sham condition itself; contrasts with true MVF will be used as supportive evidence to describe separation from MVF rather than as the primary ranking criterion. The primary deliverable of this study is therefore a prespecified ranking of candidate sham conditions and the selection of one sham condition to be carried forward into subsequent RCTs, accompanied by a frozen supplementary record (material definitions/IDs and key implementation specifications) to support reproducibility and reduce design flexibility ^[8, 17]^.

This mechanistic study is designed to generate evidence to support sham-condition selection for subsequent clinical trials, thereby improving internal validity and interpretability of MVF-related intervention effects. The protocol (v1.0) has been finalised and locked before participant recruitment, data collection, offline preprocessing, ROI-level summarisation, and any statistical analysis of condition effects. At the time of manuscript submission/posting, no participants have been enrolled and no experimental data have been collected. A subsequent version (v1.1) may include clarifications and editorial improvements to enhance reproducibility (e.g., expanded Appendix descriptions, clarified acquisition-time QC decision rules, explicit LMM specification, and fixed sham-condition naming/IDs), without changes to prespecified primary outcomes, ROIs/channel-to-ROI mapping, contrasts, multiplicity adjustment, eligibility criteria, or QC/exclusion thresholds.

The primary objective is to screen and identify the sham material condition with the most neutral fNIRS signature—that is, the condition that minimises unintended activation relative to true MVF during a single exposure. This work is designed as an acute mechanistic screening study to inform sham-condition selection for a subsequent RCT, rather than to evaluate longer-term clinical efficacy.

## Methods

### Study design

This protocol is reported in line with SPIRIT guidance for study protocols, with adaptations appropriate for an acute mechanistic crossover experiment^[11, 18, 19]^. This is a single-centre, mechanistic screening, within-participant, randomised crossover study. In a single study visit, each participant will complete four conditions (C1 true MVF and C2–C4 prespecified sham materials), with condition order counterbalanced using a Latin-square schedule.

**Figure 1.**
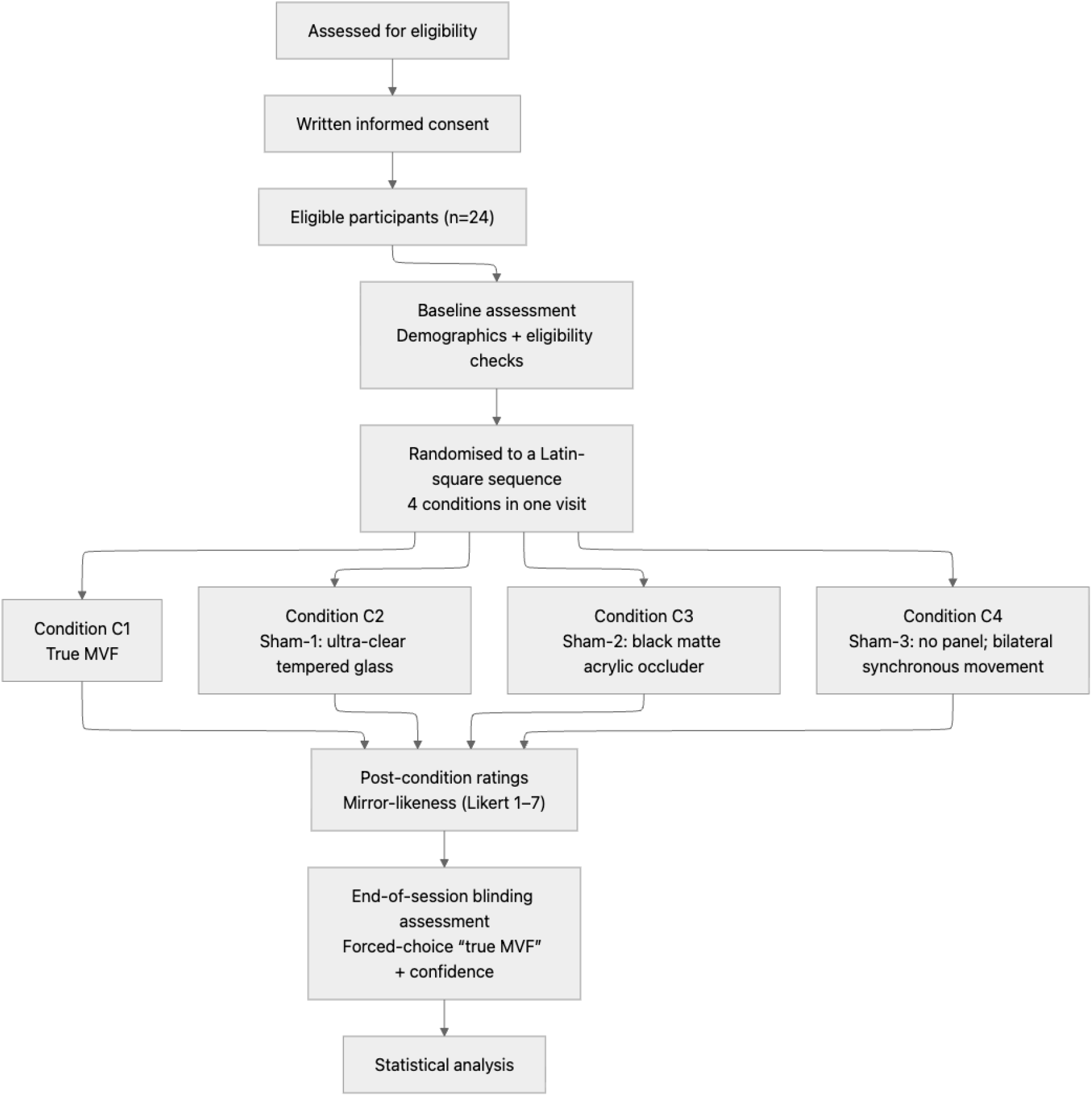
The flowchart of the trial. C1, true mirror visual feedback (MVF); C2–C4, prespecified sham material conditions; fNIRS, functional near-infrared spectroscopy.

**Table 1.**
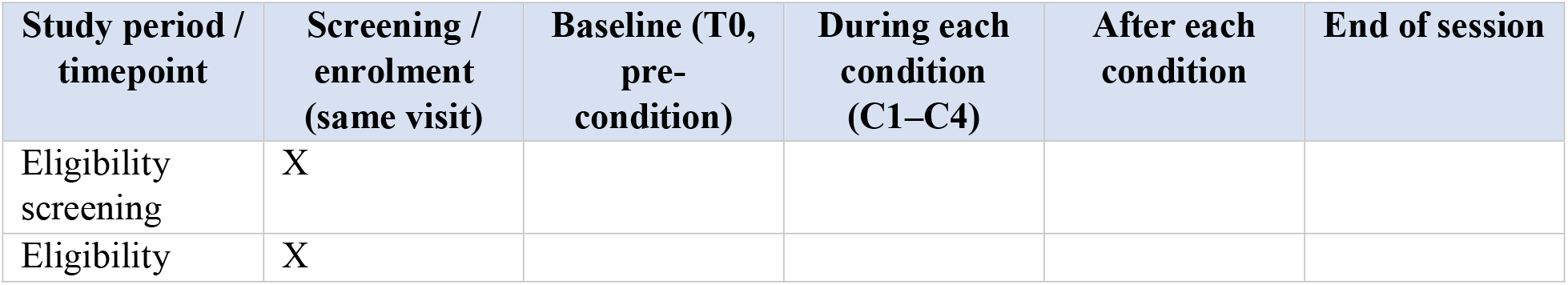
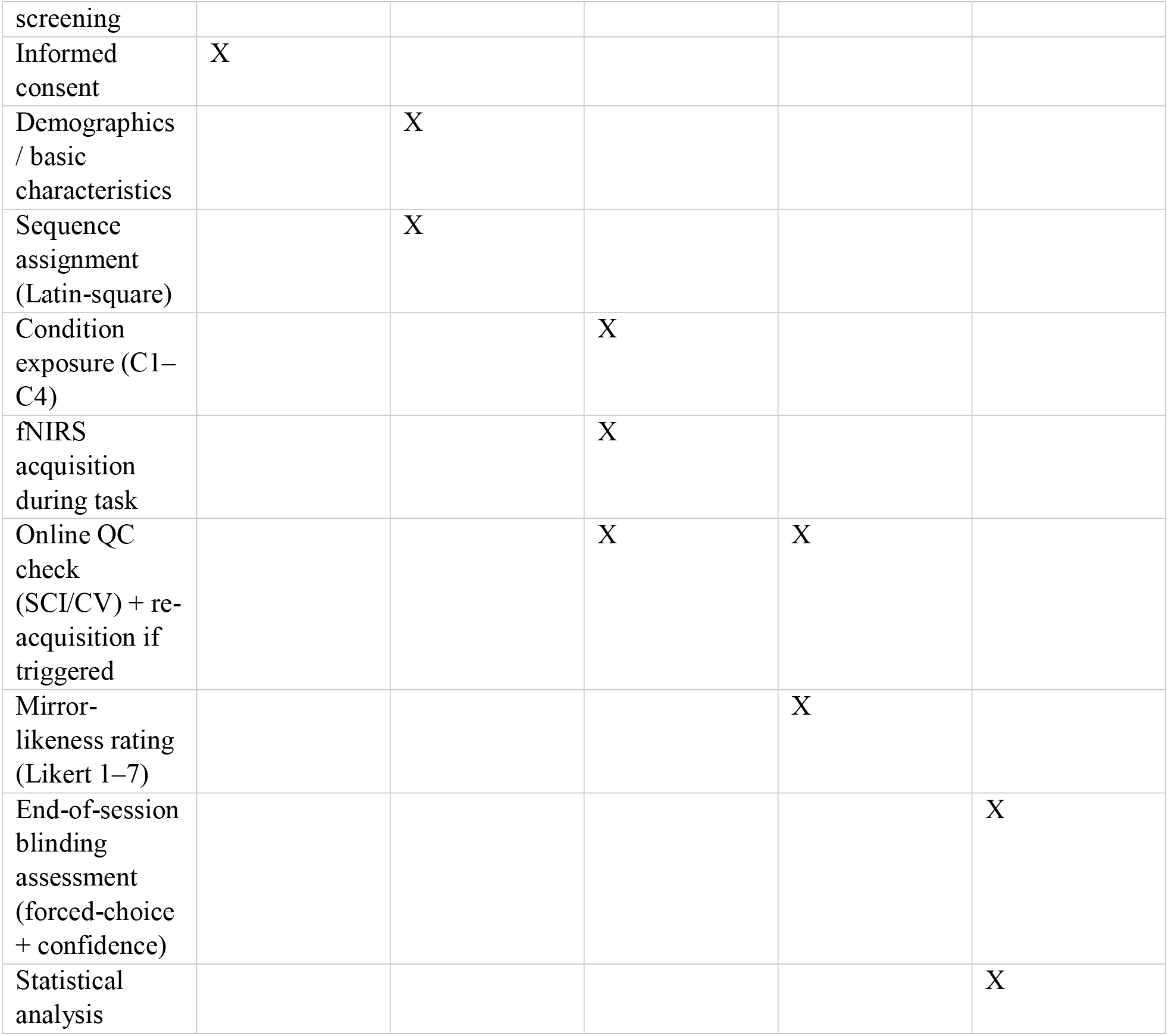
The schedule of enrolment, interventions and assessments. Abbreviations: C1, true MVF; C2–C4, sham material conditions; fNIRS, functional near-infrared spectroscopy; SCI, scalp coupling index; CV, coefficient of variation.

### Setting

The study will be conducted at Wuhan Wuchang Hospital (Rehabilitation Medicine Department), Wuhan, China, in a single standardised rehabilitation training room that also serves as the fNIRS data acquisition room.

### Participants

#### Recruitment

Participants will be recruited via WeChat-based electronic posters and referral through personal networks (word of mouth). Eligibility will be confirmed through screening (telephone prescreening and on-site assessment, as applicable). Written informed consent will be obtained prior to any study procedure.

#### Inclusion criteria

Aged 18–35 years, male or female.

Right-handed (Edinburgh Handedness Inventory score ≥60%).

No known history of neurological disorders (e.g., stroke, epilepsy, Parkinson’s disease).

No history of major chronic diseases affecting cerebral haemodynamics (e.g., uncontrolled hypertension, diabetes, severe dyslipidaemia).

Normal or corrected-to-normal vision and hearing; no colour blindness/colour weakness affecting visual tasks.

Scalp intact without lesions; able to safely wear the fNIRS cap/optodes.

Able to understand study procedures and provide written informed consent.

#### Exclusion criteria

Long-term use of centrally acting medications.

Participation in fNIRS/fMRI/EEG or mirror-therapy related experiments within the previous 3 months.

Head circumference <54 cm or >58 cm. Pregnancy or lactation.

Alcohol consumption within 24 hours, or caffeine/energy drinks within 6 hours prior to the session.

Any condition preventing completion of the full session or safe task execution, as judged by the investigator.

### Sample size

The planned sample size is **24 healthy participants**. This mechanistic, within-participant randomised crossover screening study is designed to rank candidate sham conditions based on ROI-level task-evoked fNIRS responses rather than to provide definitive hypothesis testing of clinical efficacy. Therefore, the sample size is primarily feasibility-driven^[20]^. We prespecify that **at least 20 complete analysable datasets** will be required for the primary ROI-based analyses after acquisition-time QC. Recruitment will continue until 24 participants have completed the study visit, unless the study is stopped earlier for safety, feasibility, or administrative reasons.

### Randomisation and blinding

A Latin-square sequence list will be generated in advance by a team member who is not involved in outcome analysis and stored as a locked allocation table. The experimenter will implement allocation sequentially by enrollment time, following the next available sequence on the list, and the assigned sequence ID will be recorded.

To assess the success of blinding, participants will rate perceived mirror-likeness after each condition on a 1–7 Likert scale; at the end of the session, they will be asked to identify which condition they believe was the “true MVF” condition (forced choice) and to report their confidence on a 1–7 scale.

### Interventions

Across all four conditions, the experimental context will be standardised (seating position, table height, arm placement, task instructions and timing); only the panel/material configuration will differ between conditions. Participants will be seated with the chair back at approximately 100–110°, with the apparatus positioned vertically on the table midline. A 35 cm × 45 cm planar mirror (unframed) will be used for C1; when a panel is used in sham conditions, it will match the mirror in height/width and be positioned identically. The moving (active) hand will be placed on the near side of the apparatus with the wrist crease approximately 3 cm from the apparatus edge. The contralateral hand will be positioned on the opposite side and, when required, fully occluded from view; importantly, **the contralateral hand will be instructed to remain still throughout the task in all four conditions (C1–C4)**. Thus, the grasping task will be performed by the moving (active) hand only, and the contralateral hand will serve as a stationary reference across conditions.

#### Condition 1 (C1): true mirror visual feedback (MVF)

A 35 cm × 45 cm planar mirror will be positioned vertically with the reflective surface facing the moving (active) hand. The contralateral hand will be placed behind the mirror and fully occluded from direct view. Participants will perform the standardised grasping task while viewing the mirror reflection of the moving hand, producing mirror visual feedback.

#### Condition 2 (C2): Sham-1 (ultra-clear tempered glass)

The mirror will be replaced by an ultra-clear tempered glass panel (6 mm thickness; light transmittance ≥91%), matched in height/width and positioned identically to C1. Participants will view the moving hand directly through the glass (no mirror reflection/left–right reversal).

#### Condition 3 (C3): Sham-2 (black matte acrylic occluder)

A black matte acrylic occluder (5 mm thickness; reflectance <5%) will be positioned identically to C1 and will fully block the view of both the moving and contralateral hands (no visual feedback). Participants will perform the same task under identical timing and instructions.

#### Condition 4 (C4): Sham-3 (no panel; both hands visible)

No panel (mirror/glass/occluder) will be placed. Both hands will be visible directly; however, only the moving (active) hand will perform the grasping task while the contralateral hand remains still, controlling for a no-panel visual context while removing mirror-specific visual feedback.

### Task paradigm

#### Overall structure

Each session will begin with a 6 min resting baseline. Participants will then complete four conditions. For each condition, there will be an initial 60 s rest period followed by five task blocks, each comprising 20 s rest and 20 s open-hand grasping. A fixed 3 min inter-condition interval will be provided between conditions to allow haemodynamic signals to return toward baseline, to re-check optode contact, and to minimise motion artifacts. Operationally, the first ∼30 s of this interval will be used for condition/material switching and a quick check for cable pulling/optode displacement; the remaining time is quiet rest. Data recorded during the inter-condition interval will not be included in task-evoked analyses.

#### Standardised motor task

During task blocks, participants will perform open-hand grasping without an object with the palm facing upward (supinated). Movements will be self-paced at approximately 0.5 Hz (no metronome was used). Participants will be instructed to minimise extraneous movement by keeping the non-grasping hand and the head/trunk as still as possible throughout the task.

**Figure 2.**
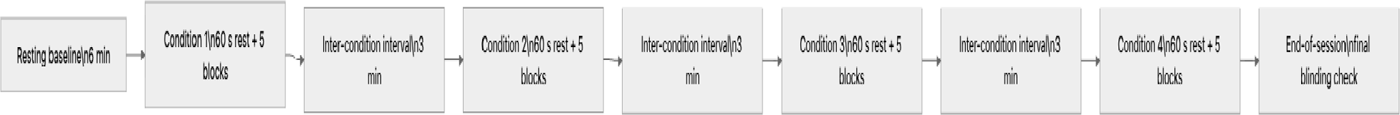
Session timeline for the within-participant crossover experiment. Following a 6 min resting baseline, each participant completes four experimental conditions (C1–C4) in a single visit. Each condition comprises a 60 s initial rest period followed by five task blocks, and consecutive conditions are separated by a fixed 3 min inter-condition interval to allow haemodynamic recovery and apparatus adjustment. The session ends with a final blinding assessment.

### Procedures and fNIRS acquisition

#### Participant timeline

The session timeline and block structure are shown above. A fixed inter-condition interval is used to allow signal recovery and to re-check optode contact; the end of session includes a final blinding check.

#### fNIRS acquisition

fNIRS data will be acquired using the NirSmart-6000A system (Danyang Huichuang Medical Equipment Co., Ltd., Jiangsu, China) with dual wavelengths (730 and 850 nm) at a sampling rate of 11 Hz. The mean source–detector distance will be 3.0 cm (range 2.7–3.3 cm). The optode array will comprise 23 sources and 15 detectors, forming 49 measurement channels. The probe montage and channel numbering are provided in Appendix A and Supplementary Figure S1 (study-specific montage with source/detector IDs and channel numbers) and will be used for the prespecified channel-to-ROI mapping.

### Quality control (acquisition-time)

Signal quality will be monitored online during acquisition. Channels will be flagged as low quality if the scalp coupling index (SCI) is <0.60 and/or the coefficient of variation (CV) is >25%. If a channel remains low quality after re-check and adjustment, it will be marked as a bad channel and excluded. If more than 20% of channels are bad for a given condition, that condition will be re-acquired once after troubleshooting; if the re-acquisition still fails, the condition will be labelled invalid. Participants will be excluded from the primary ROI analysis if fewer than two valid conditions remain. For ROI-level outcomes, an ROI×condition datapoint will be set to missing if fewer than 50% of channels mapped to that ROI remain valid after QC. Motion artifacts and physiological noise are well-recognised challenges in fNIRS and motivate prespecified QC and correction steps ^[16, 21-24]^ . Reporting of fNIRS acquisition/QC and analytic decisions will follow current best-practice recommendations for fNIRS publications ^[25]^.

### Outcomes

#### Primary outcome

The primary outcome is the ROI-level task-evoked change in oxygenated haemoglobin (ΔHbO) within prespecified ROIs (PMC and SM1/M1). Primary estimation will be based on GLM-derived β estimates, which will serve as the main inputs for inference.

#### Secondary and exploratory outcomes

As a complementary descriptive summary, ΔHbO will also be summarised within a prespecified window of 5–20 s post-onset relative to the immediately preceding rest period. Secondary and exploratory outcomes include ROI-level task-evoked changes in deoxygenated haemoglobin (ΔHbR) within the same prespecified ROIs, exploratory whole-channel analyses, and exploratory connectivity metrics, if pursued; any such connectivity analyses will be prespecified prior to analysis.

Exploratory connectivity metrics can be sensitive to thresholding and analytic choices; where connectivity is pursued, robustness checks across reasonable threshold ranges will be considered^[26]^.

#### Decision rule for sham selection (prespecified)

The “most neutral” sham material will be defined primarily as the sham condition with the smallest motor-network activation, operationalised as the smallest absolute estimated ROI-level ΔHbO within prespecified motor ROIs. Sham conditions will be ranked primarily by a composite neutrality score defined as the mean absolute ROI-level ΔHbO across bilateral SM1/M1 and PMC (four ROI×hemisphere entries). Lower scores indicate smaller absolute activation and therefore greater neural neutrality. Holm-adjusted contrasts with true MVF will be reported as supportive evidence to describe separation from MVF and to aid interpretation of MVF-likeness, but they will not constitute the primary ranking criterion. If sham conditions have closely similar composite neutrality scores, the final interpretation will consider effect sizes, 95% confidence intervals, QC robustness, and blinding/mirror-likeness ratings, without prioritising smaller absolute contrasts with true MVF.

### Data management

All data will be de-identified using study codes, with the re-identification key stored separately under restricted access. Study data will be stored securely with at least two backups. For each participant, the minimum export set will include the raw data, events/marker file, and session log (with a QC summary where feasible) to support traceability and reproducibility. Data will be retained for a minimum of 10 years. De-identified data are planned to be shared publicly 6 months after publication of the main results, subject to ethics and institutional policies.

### Monitoring plan

This is a minimal-risk, single-visit mechanistic study in healthy adults. Protocol deviations, interruptions, and QC/re-acquisition events will be documented in the session log. Any adverse events will be recorded, managed, and reported as required by the ethics committee.

### Statistical analysis plan

The data will comprise repeated measures within participants. The primary analysis will use a linear mixed-effects model with condition specified as a fixed effect and a participant-specific random intercept. A fixed period effect (1–4, corresponding to the order of conditions within the visit) will be included to account for time/ordering effects in the crossover design. Planned pairwise comparisons (six contrasts) will include C1 versus C2/C3/C4, C2 versus C3/C4, and C3 versus C4, with multiplicity controlled using the Holm procedure (two-sided α=0.05). Holm adjustment will be applied across the six planned pairwise contrasts **within each prespecified ROI** (two-sided family-wise α=0.05). Missing or invalid data will not be imputed; exclusions will follow the prespecified QC thresholds.

Analyses will be implemented using reproducible scripts and widely used fNIRS analysis toolchains where applicable (e.g., HomER and NIRS_KIT)^[27, 28]^.

## Discussion

This study addresses a key methodological challenge in trials of mirror therapy and mirror visual feedback (MVF): selecting a sham control that matches contextual features (e.g., posture, attention, task timing, and experimenter–participant interaction) while minimising MVF-specific visual input that could inadvertently engage mirror-related motor networks^[2, 5, 9]^. Using a within-participant, randomised crossover fNIRS paradigm with prespecified ROIs, contrasts, and multiplicity control, the protocol is positioned as an acute mechanistic screening experiment to identify the sham-material condition with the most neutral cortical signature during a standardised single exposure ^[16]^.

A major strength of the protocol is its emphasis on reproducibility and traceability^[17, 25]^. Core elements of the paradigm are standardised across conditions, and operational safeguards are specified, including acquisition-time QC thresholds and re-acquisition rules, standardised task timing/instructions aligned with the session log, and a frozen channel-to-ROI mapping record. In addition, the prespecified sham-selection decision rule and neutrality-ranking approach are intended to reduce analytic flexibility and support transparent interpretation of between-condition differences ^[8, 9]^.

Several limitations should be considered. First, the study is conducted in healthy adults and uses a single-session design; therefore, findings may not generalise directly to clinical stroke populations or to longer-term therapeutic effects^[20]^. Second, participant blinding may be imperfect because perceptual differences between materials can influence attention, expectancy, and engagement; blinding assessment is incorporated to quantify mirror-likeness perceptions and forced-choice identification of the true MVF condition ^[9]^. Third, fNIRS outcomes remain vulnerable to motion artifacts, variability in scalp coupling, and potential imprecision in ROI mapping^[16, 21-24]^; these risks are mitigated through prespecified preprocessing/QC procedures and documentation, but residual noise may remain.

Overall, the screening outputs from this protocol are intended to support more rigorous sham-condition selection and reporting in subsequent MVF trials^[2, 5, 9, 10]^. The resulting evidence and frozen supplementary materials will facilitate replication and downstream trial design decisions, including fidelity monitoring and transparent reporting of preprocessing and analysis choices^[25]^.

## Ethics and dissemination

Ethics approval was obtained from the Ethics Committee of Wuchang Hospital Affiliated to Wuhan University of Science and Technology (Approval No.: 2025-112-01). The study is considered minimal risk; any adverse events will be recorded, managed, and reported as required. Findings will be disseminated through publication of this protocol manuscript and a subsequent results manuscript, with key supplementary materials provided as online appendices/supplements as required by the target journal.

## Author contributions

Author contributions (CRediT): Conceptualization: Tangzhu Yang, Siyuan Wei, Yuezhu Wang, Dingqun Bai. Methodology: Tangzhu Yang, Siyuan Wei, Yuezhu Wang, Dingqun Bai. Project administration: Tangzhu Yang, Dingqun Bai. Supervision: Tangzhu Yang, Dingqun Bai. Formal analysis: Tangzhu Yang, Siyuan Wei, Yuezhu Wang. Data curation: Tangzhu Yang, Yuezhu Wang. Writing – original draft: Tangzhu Yang, Yuezhu Wang. Writing – review & editing: Tangzhu Yang, Yuezhu Wang, Dingqun Bai. Visualization: Tangzhu Yang, Yuezhu Wang. Funding acquisition: Tangzhu Yang, Dingqun Bai.

## Patient and public involvement

Patients and/or the public were not involved in the design, conduct, reporting or dissemination plans of this mechanistic screening study. This study uses a single-visit, highly standardised experimental paradigm in healthy participants to screen candidate sham mirror-therapy materials using task-evoked fNIRS outcomes; therefore, there was limited scope for meaningful patient or public co-design at this stage. Participant burden and safety considerations were addressed through ethics review and written informed consent procedures.

## Patient consent for publication

Not applicable.

## Provenance and peer review

Not commissioned; externally peer reviewed.

## Data availability statement

De-identified participant data will be made available in accordance with ethics approval and institutional policy via a public repository after publication of the primary results; access will require a methodologically sound proposal and a data use agreement prohibiting re-identification.

