## Supplementary Materials for "Screening sham mirror-therapy materials using fNIRS: protocol for an acute within-participant randomised crossover mechanistic study"

- Appendix A: Channel-to-ROI mapping and optode montage (frozen export).
- Appendix B: Sham condition material definitions/IDs (TIDieR-lite table; frozen).
- Appendix C: Preprocessing parameter freeze table.
- Appendix D: Edinburgh Handedness Inventory (EHI) details.
- Appendix E: Session log (minimum required fields).
- SPIRIT checklist.

#### Appendix A: Channel-to-ROI mapping and optode montage (frozen export).

The optode montage will be positioned according to the international 10–20 (extended) system. Channels will be assigned to ROIs a priori and **kept fixed across all analyses**.

**ROI aggregation rule:** For each condition, ROI-level ΔHbO will be computed as the **mean** of ΔHbO across **valid channels** mapped to that ROI (after QC). If fewer than **50%** of ROI-mapped channels are valid for a given ROI×condition, that ROI×condition value will be set to missing.

**Mapping source and freezing:** The channel-to-ROI mapping is **pre-specified** and will be **frozen prior to the first participant**. The mapping will be generated from the study-specific NirSmart/NirSpark montage using the built-in channel labeling and ROI/probabilistic registration outputs (with cross-checks against 10–20 anchors). The frozen mapping will be exported (channel list + montage figure), versioned, and stored in the analysis log.

**ROI assignment rule:** ROI assignment will be based on the software-generated probabilistic registration outputs (e.g., Brodmann/AAL/LPBA40 labels) anchored to 10–20 landmarks, using the default probabilistic-label rule in NirSpark/NirSpace with manual cross-checks against 10–20 anchors.

**Frozen mapping record:** The exported channel list and montage figure (including software version and export date) will be archived in the analysis log as the frozen mapping record.

**Operational definition of SM1/M1:** For ROI-based analyses, **SM1/M1 is operationalized as the union of M1 and S1 channels** (i.e., SM1 = M1 ∪ S1), consistent with the sensorimotor strip coverage of task-based fNIRS and pre-specified here to avoid post hoc ROI switching.

**Primary vs exploratory channels:** The fNIRS acquisition uses a **49-channel montage**. The 20 channels listed below constitute the **prespecified primary analysis channels** mapped to the four target ROIs (SM1/M1 and PMC, bilaterally) for the primary ROI-level analyses. The remaining **29 channels** in the 49-channel montage are not included in the primary ROI analyses; they may be used for **exploratory whole-channel analyses** only, consistent with the protocol's secondary/exploratory outcomes (if pursued).

| **ROI** | **Hemisphere** | **Approx. 10–20 anchor** | **Channels (finalised list)** |
|---|---|---|---|
| SM1/M1 | Left | C3/CP3 | CH36, CH39, CH40 |
| SM1/M1 | Right | C4/CP4 | CH27, CH28, CH30 |
| PMC | Left | FC3/F3 | CH34, CH35, CH37, CH38, CH41, CH49 |
| PMC | Right | FC4/F4 | CH13, CH14, CH25, CH26, CH29, CH48 |

**Optode montage figure:** The study-specific **49-channel montage** with **source (S) / detector (D) IDs** and **channel numbers** is provided below and is treated as the **frozen record** for channel definitions used in subsequent preprocessing/QC and ROI aggregation.

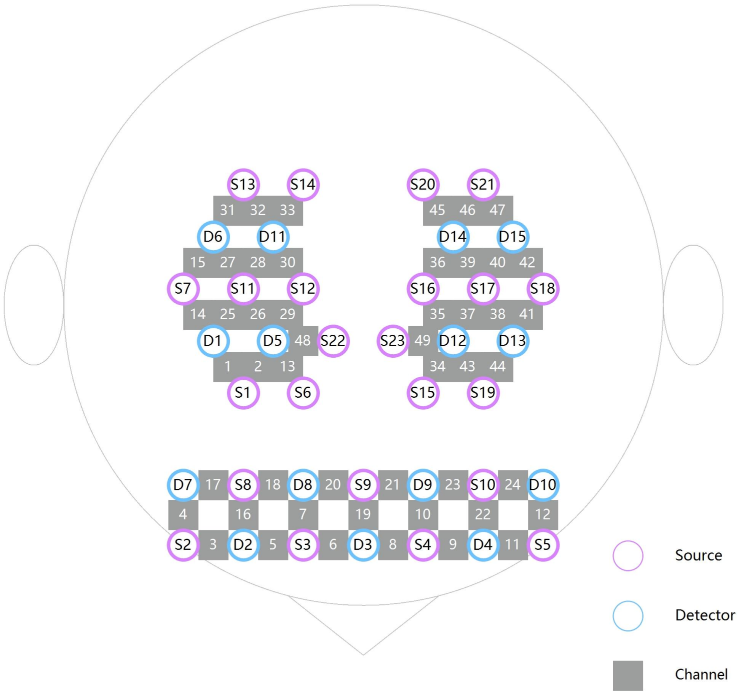

**Supplementary Figure S1. Study-specific fNIRS optode montage with channel numbering.** Purple circles denote **sources** (labelled S*); blue circles denote detectors (labelled D*); orange squares denote **measurement channels** (numbered). A channel is defined by the source–detector pair shown in the montage. This figure is treated as the frozen record for channel definitions and is used to ensure traceability of channel IDs and to support reproducible channel-to-ROI mapping (see Appendix A table). The montage was positioned according to the international 10–20 (extended) system; detailed placement and cap-fitting procedures are provided in the experimental SOP.

### Appendix B: TIDieR-lite table for sham-material conditions

This table standardizes the implementation details across the four conditions and serves as the **frozen sham material definitions/IDs record** referenced in the protocol. Material specifications available at protocol lock are summarized here; setup photos, if available, are archived locally as supporting operational records.

| **Condition** | **Material (ID/name)** | **Posture & setup** | **Visual content (participant view)** | **Executor** | **Adherence / fidelity recording** |
|---|---|---|---|---|---|
| C1: True MVF | Mirror (standard unilateral MT) | Seated; standardized table height and arm position; non-grasping hand placed behind the mirror; optodes secured; open-hand grasping without an object; movements self-paced at approximately 0.5 Hz | Mirror reflection of the moving grasping hand presented as if it were the affected/non-grasping side (classic MVF). | Trained experimenter/therapist (same executor for all sessions when feasible). | Session checklist: correct posture/hand placement; block timing adhered; major deviations (posture change, looking away, stopping) time-stamped. Video recording optional if approved. |

| Condition | Material (ID/name) | Posture & setup | Visual content (participant view) | Executor | Adherence / fidelity recording |
|---|---|---|---|---|---|
| | | (no metronome). | | | |
| C2: Sham-1 | Sham-1: ultra-clear tempered glass panel (6 mm thickness; light transmittance ≥91%; same height/width as the mirror) | Identical to C1 (same seating, table height, arm placement, and task timing). | Participants view the moving hand directly through the glass (no mirror reflection/left–right reversal), thereby minimising mirror-like visual feedback while maintaining comparable visual context. | Same executor as C1. | Session checklist + material ID logged; participant-rated mirror-likeness (1–7) recorded after each condition; executor notes any protocol deviations. |
| C3: Sham-2 | Sham-2: black matte acrylic occluder (5 mm thickness; reflectance <5%) | Identical to C1. | The occluder fully blocks the view of both hands (no visual feedback), while posture, timing, and interaction are kept identical to C1. | Same executor as C1. | Session checklist + material ID logged; deviations time-stamped; participant-rated mirror-likeness (1–7) recorded after each condition; post-session blinding questionnaire |

| Condition | Material (ID/name) | Posture & setup | Visual content (participant view) | Executor | Adherence / fidelity recording |
| --- | --- | --- | --- | --- | --- |
| | | | | | completed. |
| C4: Sham-3 | Sham-3: no panel (no mirror/glass/occluder); non-grasping hand still | Identical to C1. | Both hands are visible directly; only the moving (active) hand performs the grasping task while the contralateral (non-grasping) hand remains still, controlling for a no-panel visual context while removing mirror-specific visual feedback. | Same executor as C1. | Session checklist + material ID logged; participant-rated mirror-likeness (1–7) recorded after each condition; timing adherence verified; reasons for any deviation recorded. |

### Appendix C: Preprocessing parameter freeze table

The following preprocessing/QC parameters will be **frozen before the first participant** (v1.0) and recorded in the analysis log (NirSpark project export + analysis notebook). Any changes after freezing will require a protocol amendment and version update.

| Module | Item | Primary setting (frozen value) | Freeze time point | Recorded location |
| --- | --- | --- | --- | --- |
| Motion correction | Detection/correction method | Moving standard deviation-based detection + spline interpolation (primary) | Before first participant | Analysis log + NirSpark export |
| Filtering | Band-pass filter | 0.01–0.20 Hz (primary) | Before first participant | Analysis log + NirSpark export |

| Module | Item | Primary setting (frozen value) | Freeze time point | Recorded location |
|---|---|---|---|---|
| GLM (primary outcome) | HRF | Canonical HRF (software default; version documented) | Before first participant | Analysis log + NirSpark export |
| GLM (primary outcome) | Regressors / covariates | Condition-specific task regressor(s); additional covariates (if any) will be prespecified and listed in the analysis log before freezing | Before first participant | Analysis log |
| Quality control | SCI threshold | SCI ≥ 0.60 | Before first participant | Protocol (QC section) + analysis log |
| Quality control | CV threshold | CV ≤ 25% | Before first participant | Protocol (QC section) + analysis log |
| Outcome computation | Primary time window / baseline | 5–20 s post-onset; baseline = immediately preceding rest within each block | Before first participant | Protocol (Outcomes section) + analysis log |

### Appendix D: Edinburgh Handedness Inventory (EHI) details

Participants' handedness will be assessed using the Edinburgh Handedness Inventory (EHI). The EHI includes 10 common activities; participants indicate their preferred hand for each activity.

#### EHI items (10 everyday hand-use activities)

1. Writing
2. Drawing
3. Throwing
4. Scissors
5. Toothbrush
6. Knife (without fork)
7. Spoon
8. Broom (upper hand)

9. Striking a match
10. Opening a box lid (holding the lid)

#### Scoring and Laterality Quotient (LQ)

For each item, record whether the participant uses the **Right (R)** hand or **Left (L)** hand (optional: allow "both" only if used consistently; if "both" is allowed, it should be handled according to a prespecified rule, e.g., scored as 0.5R + 0.5L).

Compute the Laterality Quotient (LQ) as:

$$LQ = \frac{(R - L)}{(R + L)} \times 100$$

where **R** is the number of items preferentially performed with the right hand and **L** is the number of items preferentially performed with the left hand.

#### Handedness threshold for eligibility

Participants will be considered right-handed and eligible if **LQ ≥ +60**.

### Appendix E: Session log (minimum required fields)

This appendix defines the **minimum required** session-log fields to ensure standardized documentation of protocol deviations, interruptions, and data-quality issues across participants and conditions. A **printable session-log form** is provided in the corresponding *Participant Print Pack*; the items below specify the minimum content that must be captured (even if the form layout is updated in later versions).

#### Minimum required fields (per participant/session)

- **Administrative**
  - Study code
  - Session date
  - Start time / end time (or total duration)
  - Site/room (if applicable)
  - Operator/experimenter ID (initials or name)
- **Randomization / condition coding (blinding safeguard)**
  - Latin-square **sequence ID**
  - Actual execution order recorded as **conditionA–conditionD (Run1–Run4)**
  - For each Run, the corresponding allocation letter **A/B/C/D**

    - **Red line:** do **not** write any material names (e.g., mirror/glass/blackboard/empty frame) anywhere in the session log.
- **Protocol deviations / interruptions (time-stamped)**
    - Any deviation from standardized instructions (e.g., incorrect posture, looking away, non-compliance)
    - Interruptions (pause/stop), with reason and duration
    - Participant-reported discomfort or request to stop
- **fNIRS setup / QC and re-acquisition (time-stamped)**
    - Notable optode/cap adjustments (what/when/which side if applicable)
    - QC failures triggering troubleshooting (e.g., low SCI/high CV or other prespecified online QC flags), and the corrective actions taken
    - Re-acquisition events (which Run/condition, trigger reason, whether it resolved the issue)
- **Motion/physiological confounds (time-stamped)**
    - Coughing, speaking, visible head/trunk movement
    - Any event judged to compromise block structure or data validity (brief description)
- **File/traceability notes**
    - Any file-naming exceptions or missing exports (raw/events/QC summaries) and how they were resolved
    - Software/hardware version notes if changed from standard setup

#### Recording rules

- All major events should be **time-stamped** and should match corresponding event markers where applicable (e.g., the events/marker file).
- Use the session log to document **what happened**, **when it happened**, and **whether it may affect** a specific Run/block; avoid interpretation beyond factual notes.
